# Longitudinal Investigation of Autonomic Symptoms in Biologically Confirmed (SAA+) Parkinson’s Disease

**DOI:** 10.64898/2026.11.09.26367184

**Authors:** Adeel A. Memon, Seung Ho Choi, Jie Yin, David-Erick Lafontant, Chelsea Caspell-Garcia, Daniel Weintraub, Amy W. Amara, Marina Picillo, The Parkinson’s Progressive Marker Initiative

## Abstract

**Background:** Autonomic symptoms are common in Parkinson’s disease (PD) and impact quality of life, with unclear relationship to motor and cognitive progression.

**Objective:** Characterize five-year progression of autonomic symptoms in early PD and their association with motor and cognitive outcomes.

**Methods:** We analyzed data from 874 PD participants and 252 healthy controls (HC) enrolled in Parkinson’s Precision Medicine Initiative (PPMI). Autonomic symptoms were assessed annually over five years using Scales for Outcomes in Parkinson’s disease-Autonomic dysfunction (SCOPA-AUT); affected autonomic domains were defined as domain subscore >1. Clinical measures included MDS-UPDRS and Montreal Cognitive Assessment. Generalized estimating equations evaluated longitudinal changes, adjusting for age, sex, medication, and comorbidities.

**Results:** At baseline, 71% of PD participants reported >1 affected autonomic domain, increasing to 85% among 287 PD participants with 5-year follow-up; HCs reporting >1 affected autonomic domain increased from 40% of participants at baseline to 46% among 145 participants with 5-year follow-up. Gastrointestinal and urinary symptoms were most prevalent in PD. Medication use for constipation and urinary dysfunction increased over time in PD. Cardiovascular comorbidities were common and increased in prevalence in both groups. Greater baseline autonomic symptom burden predicted more rapid cognitive, but not motor decline in PD (p=0.002).

**Conclusion:** Autonomic symptoms in PD are common at disease onset, near universal after 5 years, and associated with faster cognitive, but not motor, decline, driven by those with severe symptoms. The high prevalence and impact of autonomic symptoms in early PD highlights the importance of early assessment and management of these symptoms.

**Ethical Compliance Statement:** All study and recruitment materials were approved by institutional review boards or ethics committees at each site. Written informed consent was obtained from all participants before undergoing any study evaluations. The study was performed in accordance with the principles outlined in the Declaration of Helsinki and with Good Clinical Practice guidelines. We confirm that we have read the Journal’s position on issues involved in ethical publication and affirm that this work is consistent with those guidelines.

**Funding Disclosures:** PPMI – a public-private partnership – is funded by the Michael J. Fox Foundation for Parkinson’s Research and funding partners, including AbbVie, Alamar Biosciences, Aligning Science Across Parkinson’s (ASAP), Arrowhead Pharma, Arvinas, AskBio, BIAL, BioArctic, Biohaven, BlueRock Therapeutics, Bristol Myers Squibb, Calico Labs, Capsida Biotherapeutics, Critical Path Institute, DaCapo Brainscience, Denali, Edmond J. Safra Foundation, Eli Lilly, Gain Therapeutics, GE Healthcare, Genentech, GSK, Insitro, Johnson & Johnson Innovative Medicine, Lundbeck, Merck, Neumora, Neuron23, Novartis, Olink, Regeneron, Roche, Sanofi, Tenvie, UCB, Vanqua Bio, Voyager Therapeutics, The Weston Family Foundation.

## Introduction

Parkinson’s disease (PD) is a progressive neurodegenerative disorder classically defined by motor symptoms of bradykinesia, rigidity, resting tremor, and postural instability[1]. However, PD is increasingly recognized as a multisystem condition with diverse non-motor symptoms that profoundly impact quality of life[1]. Among these, autonomic dysfunction, affecting cardiovascular, gastrointestinal, urogenital, thermoregulatory, and pupillomotor functions, is particularly prevalent and debilitating[2]. Studies estimate that 70–80% of PD patients experience autonomic symptoms, which can emerge early in the disease course, even preceding motor manifestations, and may predict faster progression[1, 3]. Despite its clinical significance, autonomic dysfunction remains understudied, with gaps in longitudinal data on symptom progression, pathophysiological mechanisms, and its relationship to motor and cognitive decline[3].

The pathophysiology of autonomic symptoms in PD involves both central and peripheral nervous system degeneration. Alpha-synuclein aggregates, a hallmark of PD pathology, are found in autonomic nuclei and peripheral nerves[4], suggesting that autonomic pathways may serve as conduits for the spread of pathology from the gut or other peripheral sites to the brain[5]. In PD, cardiovascular autonomic dysfunction is driven primarily by postganglionic sympathetic denervation, which impairs baroreflex function and reduces norepinephrine release from sympathetic nerve terminals, resulting in neurogenic orthostatic hypotension and inadequate vasoconstriction on standing[6]. Similarly, gastrointestinal disturbances like constipation are linked to enteric nervous system involvement, while urinary symptoms like urinary incontinence, urinary retention, voiding dysfunction, urgency and frequency stem from degeneration of Onuf’s nucleus in the spinal cord[5]. These disruptions not only impair daily functioning but also exacerbate other non-motor symptoms, such as sleep disturbances and depression, further diminishing quality of life[1].

Existing research highlights critical gaps. First, while cross-sectional studies document autonomic symptom prevalence, longitudinal data are sparse, limiting insights into temporal progression and its interaction with disease milestones[3, 7–10]. Second, the contribution of specific comorbidities, such as cardiovascular and metabolic disorders, to autonomic symptoms remains poorly quantified, potentially confounding observed associations[11]. Third, the links between autonomic impairment and motor and cognitive decline are unclear[9].

This study addresses these gaps by analyzing longitudinal data from the Parkinson’s Precision Medicine Initiative (PPMI) cohort to track autonomic symptom progression over 5 years and its relationship to motor and cognitive trajectories. Using the Scales for Outcomes in Parkinson’s disease-Autonomic Dysfunction (SCOPA-AUT)[12], we evaluate rates of change across six autonomic domains while adjusting for comorbidities and medication effects, providing a nuanced understanding of autonomic dysfunction’s natural history. Additionally, by correlating autonomic severity with dopaminergic imaging markers (i.e., DaTscan uptake in the striatum), we explore potential biomarkers for disease progression. We also incorporate orthostatic vital signs as an objective measure of autonomic function. These findings aim to inform early intervention strategies and personalized management approaches that can ultimately improve quality of life for individuals living with PD.

## Methods

### Study Population

This study analyzes data from PD and healthy control (HC) participants enrolled in PPMI[13], a large, multicenter observational study designed to identify biomarkers of PD progression. At study entry, PD participants had early-stage, idiopathic PD, enrolled within two years of diagnosis, were not on medication for treatment of PD, had a dopaminergic deficit confirmed by imaging, and did not exhibit atypical parkinsonian syndromes[13]. Additionally, individuals enrolled in the PD cohort who had a negative cerebrospinal fluid (CSF) alpha-synuclein seed amplification assay (SAA) were excluded from this analysis. HC participants were age- and sex-matched to the PD group, had no neurological disorders, and had no first-degree family members with PD[13]. Additionally, HC participants with a positive CSF SAA were excluded from this analysis. All PPMI participants provided written informed consent according to the study protocol and applicable ethical regulations[13].

Our analysis included PPMI participants with autonomic function data up to five years of follow-up. Longitudinal analyses were limited to participants with at least one annual follow-up due to adjustment for baseline response. This secondary analysis of de-identified PPMI data was conducted following PPMI data usage guidelines and did not require further ethical review.

Data used in the preparation of this article were obtained on January 8, 2024 from the Parkinson’s Precision Medicine Initiative (PPMI) database (https://www.ppmi-info.org/access-data-specimens/download-data), RRID:SCR_006431. For up-to-date information on the study, visit https://www.ppmi-info.org. This analysis was conducted by the PPMI Statistics Core and used actual dates of activity for participants, a restricted data element not available to public users of PPMI data. Statistical analysis codes used to perform the analyses in this article are shared on Zenodo (https://doi.org/10.5281/zenodo.15739155).

### Clinical Assessments and Demographics

At baseline, age, sex, age at PD symptom onset, disease duration, Movement Disorder Society-Unified Parkinson’s Disease Rating Scale parts I-III (MDS-UPDRS I-III), orthostatic vital signs (considered positive if ≥20 mm Hg change in systolic BP or ≥10 mm Hg change in diastolic BP; measured supine to standing within 3 minutes), and Hoehn and Yahr stage were recorded. Disease duration was calculated from the reported date of PD diagnosis to the baseline visit.

The MDS-UPDRS I-III was administered at each visit to assess motor symptom severity and progression, including tremor, rigidity, bradykinesia, and postural instability. The SCOPA-AUT questionnaire[12] evaluated autonomic symptoms across six domains: gastrointestinal, urinary, cardiovascular, thermoregulatory, pupillomotor, and sexual. The Montreal Cognitive Assessment (MoCA)[14] screened for cognitive impairment. Detailed information on medication usage, including those prescribed for autonomic dysfunction or other comorbidities, was collected at each visit. Data on the presence of comorbidities, such as cardiovascular, metabolic, and urinary conditions, were also gathered (Supplementary Table 4 for full list). This information allowed for adjustments in statistical analyses to account for the potential influence of comorbidities on study outcomes and allowed for a comprehensive analysis of the relationship between autonomic dysfunction, motor symptoms, cognitive decline, and other clinical outcomes in PD.

### Statistical Analysis

Data were examined for autonomic symptoms (as measured on the SCOPA-AUT) and clinical measures from baseline to year 5 where SCOPA-AUT data were present. Two distinct SCOPA-AUT metrics are reported. First, an affected autonomic domain was defined as a SCOPA-AUT subscore >1 within one of the six domains. The number of affected autonomic domains was calculated by summing across the six domains, yielding a score ranging from 0 to 6. Autonomic symptom severity was then categorized as none (0 affected domains), mild (1–2 affected domains), or severe (>2 affected domains). Second, any autonomic symptom was defined as SCOPA-AUT total score >1 summed across all items. Continuous SCOPA-AUT scores were considered but not used to emphasize domain prevalence.

Frequency (percentage) is reported for categorical measures and mean (standard deviation) or median (IQR) for continuous measures. For selected comparisons at baseline between cohorts (PD and HC), the chi-square test is presented for categorical measures, and Student’s t-test for continuous measures.

To analyze the longitudinal progression of autonomic symptoms, generalized estimating equations (GEEs) with a logit link were applied to evaluate whether the cohorts differed in their longitudinal trajectories under the assumption that the log odds of the response changed linearly over time. Specifically, cohort, time, and their interaction were included while adjusting for age, sex, and baseline response. As appropriate, models were also adjusted for medication use (yes/no) and comorbidities (yes/no).

GEEs were also used to evaluate whether the longitudinal trajectory of clinical outcomes differed by autonomic symptom severity at baseline (categorized as none (0 affected domains), mild (1–2 affected domains), or severe (>2 affected domains).) within PD participants, assuming that the clinical outcome changed linearly over time. Baseline symptom severity, time, and their interaction were included while adjusting for age, sex, baseline response. Models were also adjusted for medication use (yes/no) and comorbidities (yes/no). For MDS-UPDRS III (ON), the model also adjusted for levodopa equivalent dose (LED).

In both sets of models, the auto regressive working correlation structure was employed. Wald tests were used to assess if the interaction term between group (i.e., cohort or baseline symptom severity) and time was statistically significant. Time effect p-values were reported for all models, and separate time effects with 95% confidence intervals (95% CI) in each group were also provided when the interaction term was significant. To account for multiple comparisons, a Bonferroni corrected p-value (0.05 divided by the total number of comparisons in each table) was used for each group of outcomes, i.e., 0.05/10 = 0.005 for presence of autonomic symptoms and 0.05/7 = 0.007 for severity of autonomic symptoms in PD. Although the Bonferroni method is very conservative, it was selected for simplicity. Since this analysis is purely observational, interpretation of significant p-values prior to the Bonferroni adjustments (i.e., using the 0.05 significance level) are provided and may provide some clinically meaningful insight.

Sensitivity analyses were conducted to assess whether differential availability of follow-up autonomic data may have influenced the study findings. Baseline characteristics were compared between participants with PD with and without observed Year 5 autonomic data. In order to evaluate the potential impact of informative drop-out on the results of our longitudinal models, we incorporated inverse probability weighting (IPW) into the generalized estimating equation (GEE) models evaluating longitudinal changes in clinical outcomes by baseline autonomic symptom severity groups in the PD cohort.

IPWs were calculated as the inverse of the estimated cumulative probability of having observed autonomic data at each follow-up visit. These probabilities were estimated using visit-specific logistic regression models of the probability of being observed at a given visit conditional on baseline characteristics (age, sex, disease duration, and lowest putamen ratio) and prior-visit clinical outcomes (MDS-UPDRS Parts I, II, and III (ON) scores; Montreal Cognitive Assessment (MoCA) score; and changes in systolic and diastolic blood pressure. Clinical outcomes with intermittent missing values were imputed using linear interpolation. Values outside the observed range were not extrapolated. Participants with characteristics associated with a lower predicted probability of remaining observed received greater weight.

The resulting visit-level IPWs were applied in longitudinal GEE models detailed above.

All analyses were performed using SAS 9.4 (SAS/STAT 15.2; SAS Institute Inc., Cary, NC, USA).

## Results

### Demographics and Baseline Characteristics

Table 1 summarizes the demographic characteristics of the study cohorts. The PD cohort (N=874) was slightly older, with a mean age of 63.1 years (SD = 9.4), while the HC cohort (N=252) had a mean age of 61.3 years (SD = 11.7). The sex distribution was similar between groups, with males comprising 66% of the PD cohort and 62% of the HC cohort. These baseline characteristics were accounted for in subsequent statistical adjustments. Table 1 also shows the baseline disease characteristics for PD participants.

**Table 1:** Baseline Demographics and Clinical Characteristics.

| Variable | PD (N=874) | HC (N=252) | *p*-value |
|---|---|---|---|
| **Age at enrollment, years,** mean (SD) | 63.1 (9.4) | 61.3 (11.7) | 0.03 |
| Min.-Max. | 33.7-84.9 | 30.4-85.3 |  |
| **Male sex**, n (%) | 573 (66%) | 157 (62%) | 0.34 |
| **Age at PD symptom onset**, mean (SD) | 60.8 (9.7) | - | - |
| Min.-Max. | 25.4-83.7 | - |  |
| **Disease duration at enrollment**, median (IQR) | 0.5 (0.3-1.0) | - | - |
| Min.-Max. | 0.0-3.6 | - |  |
| **MDS-UPDRS III (OFF) score**, mean (SD) | 22.4 (9.7) | - | - |
| Min.-Max. | 3.0-62.0 | - |  |
| **Hoehn & Yahr stage (OFF)**, n (%) |  | - | - |
| 1 | 316 (36%) | - |  |
| 2 | 553 (63%) | - | - |
| 3 | 5 (1%) |  |  |

Note: Known SAA- PDs and SAA+ HCs were excluded at the time of the data download.
Age at enrollment was compared using Student’s *t*-test, and sex was compared using the chi-square test.

### Frequency of Affected Autonomic Domains

Figure 1 shows the percentage of participants reporting autonomic symptoms in each cohort over 5 years. Supplementary Table 1 presents the frequency of number of affected autonomic domains reported longitudinally over 5 years in both cohorts. At baseline, 71% of PD participants reported experiencing more than one affected autonomic domain, compared with only 40% in HC participants. Over the five-year follow-up, the proportion of PD participants with more than one affected autonomic domain increased to 85%, whereas the HC also increased, but remained lower compared to PD at 46%.

**Figure 1:**
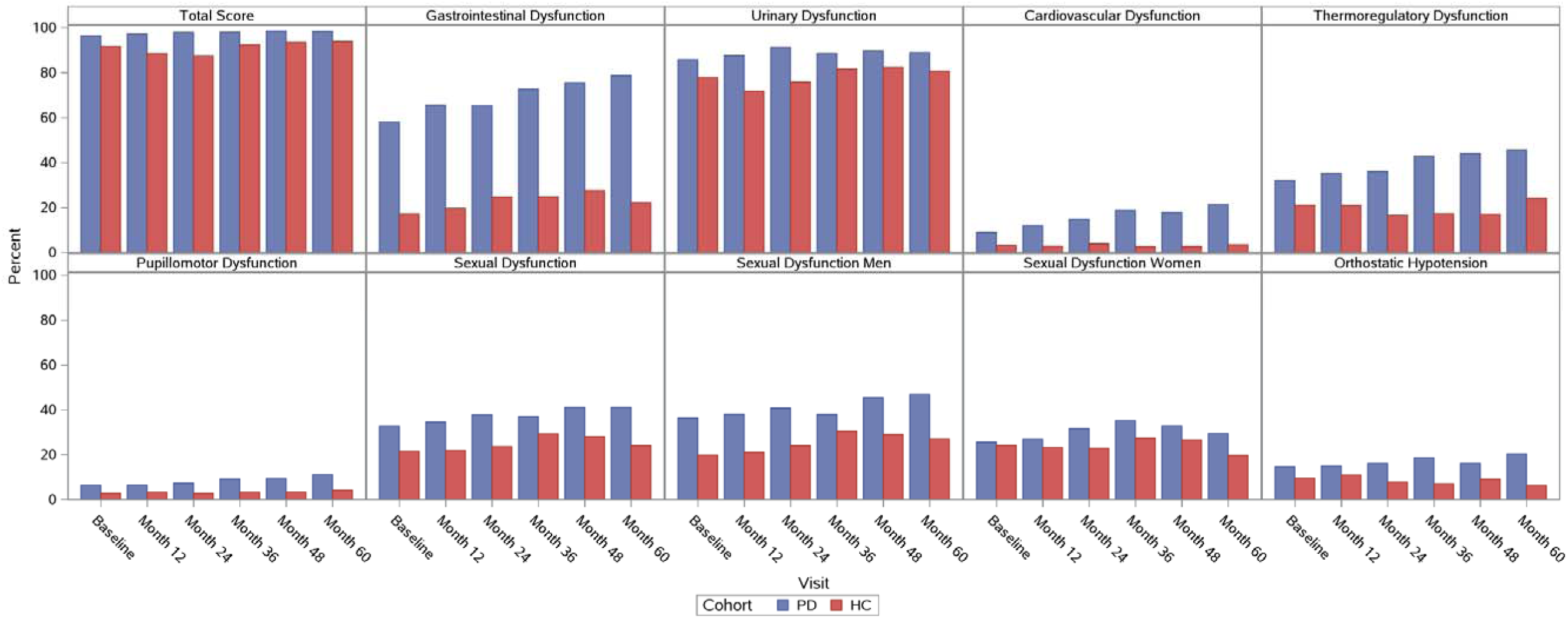
Percentage of Participants with Autonomic Symptoms and Affected Domains Over Time. Percentage of participants scoring >1 on SCOPA-AUT total,each SCOPA-AUT subscore, and experiencing orthostatic hypotension are on the y-axis and time is on the x-axis.

### Medications for Autonomic Dysfunction

Supplementary Table 2 provides an overview of medications taken for autonomic dysfunction. At baseline, 17% of PD participants were using medication for constipation, increasing to 34% by year five. Similarly, the percentage of PD participants taking medication for urinary dysfunction rose from 12% to 19%. The use of medications for sexual dysfunction remained relatively stable in both cohorts, while no HC participants reported taking medication for hypotension at any time point. These trends suggest an increasing need for pharmacological management of autonomic dysfunction in PD over time.

### Comorbidities

Supplementary Table 3 details the prevalence of comorbidities in both groups. Cardiovascular comorbidities were the most prevalent, affecting 38% of PD participants at baseline and increasing to 53% by year five. In contrast, metabolic and urinary comorbidities showed smaller increases over time. The HC group exhibited a similar pattern, but with urinary comorbidities at a consistently lower prevalence compared to PD. These findings underscore the progressive nature and more extensive involvement of autonomic systems in PD.

### Logistic Models for Autonomic Dysfunction

Table 2 presents the progression of autonomic symptoms over time in both PD and HC cohorts exhibiting SCOPA-AUT scores >1. Supplementary Table 4 shows the model adjustments for Table 2.

**Table 2:** Autonomic Symptoms and Affected Domains Over Time in PD and HC.

| **Cohort<br>PD<br>HC** | **Baseline<br>N=874<br>N=252** | **Month 12<br>N=571<br>N=219** | **Month 24<br>N=408<br>N=182** | **Month 36<br>N=325<br>N=157** | **Month 48<br>N=309<br>N=153** | **Month 60<br>N=287<br>N=145** | **p-value<br>Interaction<br>(Cohort x Time)** | **p-value<br>Time** | **OR (95% CI)** |
|---|---|---|---|---|---|---|---|---|---|
| **SCOPA AUT Total Score (total score > 1)** |  |  |  |  |  |  |  |  |  |
| PD | 842 (96%) | 555 (97%) | 399 (98%) | 318 (98%) | 304 (98%) | 282 (98%) | 0.422 | 0.012 | 1.173 (1.036, 1.329) |
| HC | 231 (92%) | 194 (89%) | 159 (87%) | 145 (92%) | 143 (93%) | 136 (94%) |  |  |  |
| **Gastrointestinal Dysfunction (subscore > 1)** |  |  |  |  |  |  |  |  |  |
| PD | 507 (58%) | 374 (65%) | 266 (65%) | 236 (73%) | 233 (75%) | 226 (79%) | 0.011 | PD: <.001<br>HC: 0.503 | PD: 1.241 (1.146, 1.344)<br>HC: 1.039 (0.929, 1.162) |
| HC | 43 (17%) | 43 (20%) | 45 (25%) | 39 (25%) | 42 (27%) | 32 (22%) |  |  |  |
| **Urinary Dysfunction (subscore > 1)** |  |  |  |  |  |  |  |  |  |
| PD | 749 (86%) | 500 (88%) | 372 (91%) | 288 (89%) | 277 (90%) | 255 (89%) | 0.104 | 0.001 | 1.138 (1.053, 1.231) |
| HC | 196 (78%) | 157 (72%) | 138 (76%) | 128 (82%) | 126 (82%) | 117 (81%) |  |  |  |
| **Cardiovascular Dysfunction (subscore > 1)** |  |  |  |  |  |  |  |  |  |
| PD | 78 (9%) | 67 (12%) | 60 (15%) | 61 (19%) | 55 (18%) | 61 (21%) | 0.297 | 0.062 | 1.129 (0.994, 1.283) |
| HC | 8 (3%) | 6 (3%) | 7 (4%) | 4 (3%) | 4 (3%) | 5 (3%) |  |  |  |
| **Thermoregulatory Dysfunction (subscore > 1)** |  |  |  |  |  |  |  |  |  |
| PD | 280 (32%) | 201 (35%) | 147 (36%) | 139 (43%) | 136 (44%) | 131 (46%) | 0.088 | 0.025 | 1.087 (1.011, 1.170) |
| HC | 53 (21%) | 46 (21%) | 30 (16%) | 27 (17%) | 26 (17%) | 35 (24%) |  |  |  |
| **Pupillomotor Dysfunction (subscore > 1)** |  |  |  |  |  |  |  |  |  |
| PD | 56 (6%) | 37 (6%) | 30 (7%) | 30 (9%) | 29 (9%) | 32 (11%) | 0.636 | 0.051 | 1.160 (0.999, 1.347) |
| HC | 7 (3%) | 7 (3%) | 5 (3%) | 5 (3%) | 5 (3%) | 6 (4%) |  |  |  |
| **Sexual Dysfunction (subscore > 1)** |  |  |  |  |  |  |  |  |  |
| PD | 285 (33%) | 197 (35%) | 154 (38%) | 120 (37%) | 127 (41%) | 118 (41%) | 0.623 | 0.017 | 1.088 (1.015, 1.166) |
| HC | 54 (21%) | 48 (22%) | 43 (24%) | 46 (29%) | 43 (28%) | 35 (24%) |  |  |  |
| **Sexual Dysfunction Men (subscore > 1)** |  |  |  |  |  |  |  |  |  |
| PD | 208 (36%) | 146 (38%) | 112 (41%) | 81 (38%) | 93 (45%) | 90 (47%) | 0.969 | 0.004 | 1.135 (1.042, 1.237) |
| HC | 31 (20%) | 28 (21%) | 27 (24%) | 29 (31%) | 27 (29%) | 24 (27%) |  |  |  |
| **Sexual Dysfunction Women (subscore > 1)** |  |  |  |  |  |  |  |  |  |
| PD | 77 (26%) | 50 (27%) | 42 (32%) | 39 (35%) | 34 (33%) | 28 (29%) | 0.555 | 0.879 | 1.010 (0.892, 1.142) |
| HC | 23 (24%) | 20 (23%) | 16 (23%) | 17 (27%) | 16 (27%) | 11 (20%) |  |  |  |
| **Orthostatic Hypotension (≥ 20 mm Hg change in systolic BP** *or* **≥ 10 mm Hg change in diastolic BP)** |  |  |  |  |  |  |  |  |  |
| PD | 126 (14%) | 83 (15%) | 64 (16%) | 60 (19%) | 49 (16%) | 58 (20%) | 0.054 | 0.790 | 0.985 (0.883, 1.099) |
| HC | 24 (10%) | 24 (11%) | 14 (8%) | 11 (7%) | 14 (9%) | 9 (6%) |  |  |  |

Note: All models are adjusted for age at baseline, sex, and baseline response.  Each model also adjusts for appropriate medication use and comorbidities shown in Appendix 1.
Descriptive summaries (mean [SD] or n [%]) included 874 participants with PD and 252 healthy controls (HCs). The longitudinal analysis included 589 participants with PD and 223 HCs who had autonomic data at baseline and at least one follow-up visit and assessed the longitudinal progression of autonomic symptoms from Year 1 through Year 5.
Significance level for comparisons is p<0.005 (after Bonferroni correction).

#### Overall Autonomic Dysfunction

The prevalence of at least mild autonomic symptoms on the SCOPA-AUT (score >1) remained consistently higher in PD participants compared to HC and was high in both groups; however, there was no significant difference in the odds of experiencing any autonomic dysfunction between the PD and HC cohorts over time (p = 0.422). Both groups exhibited significant increases over time, such that every year, the estimated odds of experiencing any autonomic dysfunction increased (p = 0.012) by a multiplicative factor of 1.173 (95% CI: 1.036, 1.329).

#### Gastrointestinal Dysfunction

PD participants had higher reporting of GI complaints compared to HC and the odds of experiencing gastrointestinal dysfunction (subscore > 1) changed differently over time between PD and HC cohorts (p = 0.011). Every year, the estimated odds of experiencing gastrointestinal dysfunction in the PD cohort increased by a multiplicative factor of 1.241 (95% CI: 1.146, 1.344; p < 0.001) compared with the HC participants whose odds did not significantly change (p = 0.503). After adjustment for multiple comparisons, the interaction is no longer significant.

#### Urinary Dysfunction

There was no significant difference in the odds of experiencing urinary dysfunction between PD and HC participants over time (p = 0.104). Every year, the estimated odds of experiencing urinary dysfunction increased (p = 0.001) by a multiplicative factor of 1.138 (95% CI: 1.053, 1.231).

#### Cardiovascular Dysfunction

The odds of cardiovascular dysfunction did not differ significantly between PD and HC participants (p = 0.297). Similarly, the odds of cardiovascular dysfunction did not change significantly with each additional year (p = 0.062).

#### Thermoregulatory Dysfunction

There was no significant difference in the odds of experiencing thermoregulatory dysfunction between PD and HC participants over time (p = 0.088). Every year, the estimated odds of experiencing thermoregulatory dysfunction increased (p = 0.025) by a multiplicative factor of 1.087 (95% CI: 1.011, 1.170).

#### Pupillomotor Dysfunction

There was no significant difference in the odds of experiencing pupillomotor dysfunction between PD and HC participants (p = 0.636). Every year, the odds of experiencing pupillomotor dysfunction did not significantly change (p = 0.051).

#### Sexual Dysfunction

There was no significant difference in the odds of experiencing sexual dysfunction between PD and HC participants over time (p = 0.623). Every year, the estimated odds of experiencing sexual dysfunction changed (p = 0.017) by a multiplicative factor of 1.088 (95% CI: 1.015, 1.166). This difference in odds appears to be more elevated in men where the odds of experiencing sexual dysfunction increased every year (p = 0.004) by a multiplicative factor of 1.135 (95% CI: 1.042, 1.237), compared to women who did not significantly change (p = 0.879) over time.

#### Orthostatic Hypotension

There was no significant difference in the odds of experiencing orthostatic hypotension between PD and HC participants over time (p = 0.054), and every year, the estimated odds of experiencing orthostatic hypotension did not significantly change (p = 0.790).

### Effect of Autonomic Symptoms on Clinical Outcomes

Table 3 examines the selected clinical measures in the PD cohort stratified by baseline autonomic symptom severity (defined by the number of symptoms present: 0 [none], 1-2 [mild], >2 [severe]) at each time point. Supplementary Table 5 shows the model adjustments for Table 3.

**Table 3:**
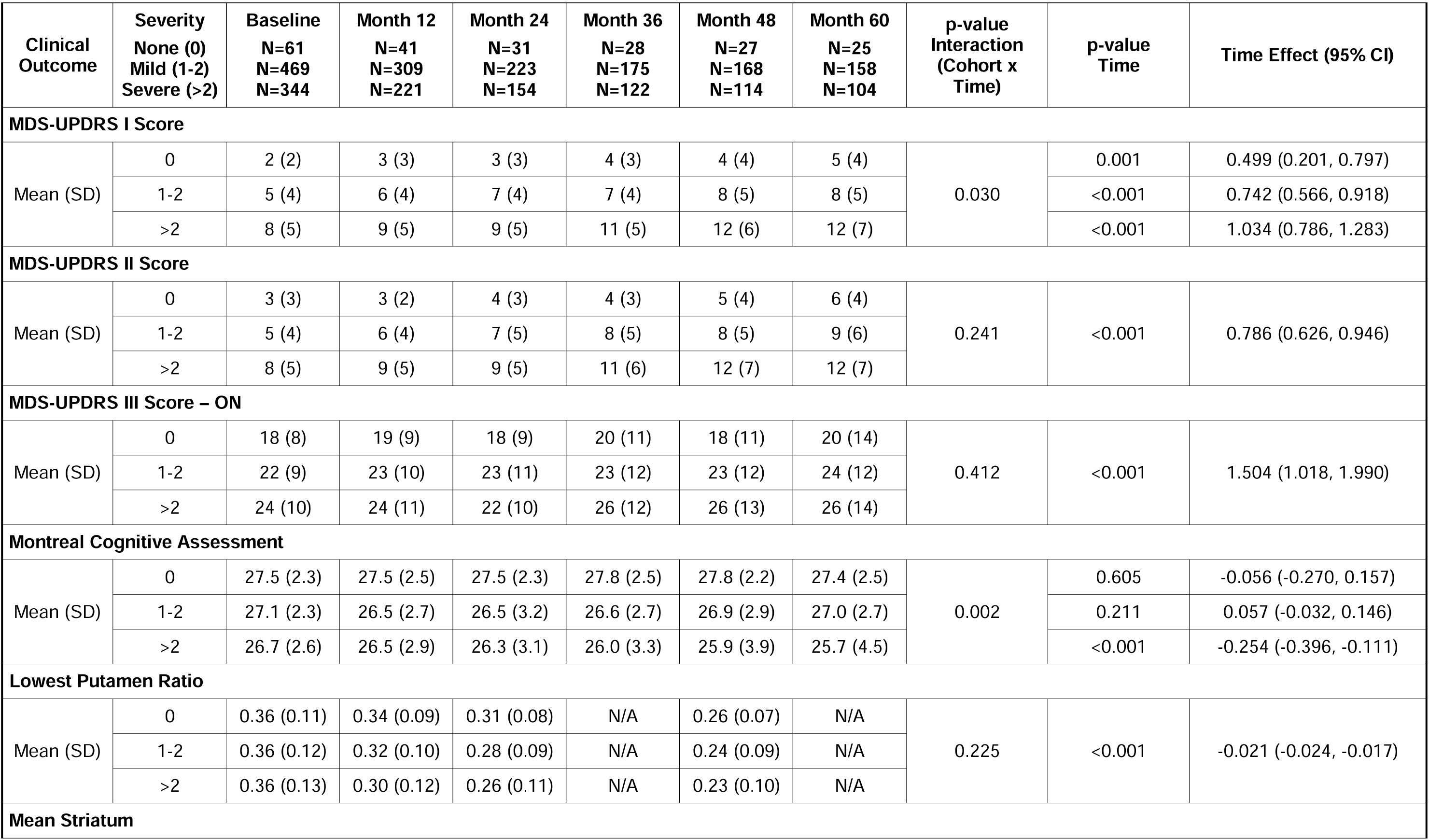
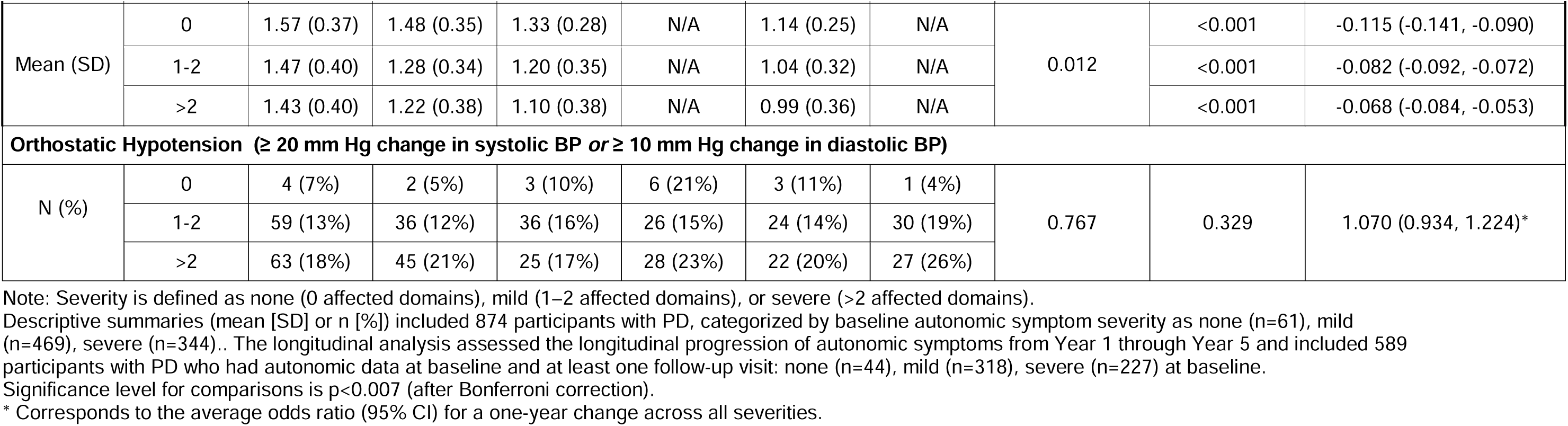
Effect of Autonomic Symptom Severity at Baseline on Clinical Outcomes in PD.

| Clinical Outcome | Severity<br>None (0)<br>Mild (1-2)<br>Severe (>2) | Baseline<br>N=61<br>N=469<br>N=344 | Month 12<br>N=41<br>N=309<br>N=221 | Month 24<br>N=31<br>N=223<br>N=154 | Month 36<br>N=28<br>N=175<br>N=122 | Month 48<br>N=27<br>N=168<br>N=114 | Month 60<br>N=25<br>N=158<br>N=104 | p-value Interaction (Cohort x Time) | p-value Time | Time Effect (95% CI) |
|---|---|---|---|---|---|---|---|---|---|---|
| **MDS-UPDRS I Score** |  |  |  |  |  |  |  |  |  |  |
| Mean (SD) | 0 | 2 (2) | 3 (3) | 3 (3) | 4 (3) | 4 (4) | 5 (4) | 0.030 | 0.001 | 0.499 (0.201, 0.797) |
|  | 1-2 | 5 (4) | 6 (4) | 7 (4) | 7 (4) | 8 (5) | 8 (5) |  | <0.001 | 0.742 (0.566, 0.918) |
|  | >2 | 8 (5) | 9 (5) | 9 (5) | 11 (5) | 12 (6) | 12 (7) |  | <0.001 | 1.034 (0.786, 1.283) |
| **MDS-UPDRS II Score** |  |  |  |  |  |  |  |  |  |  |
| Mean (SD) | 0 | 3 (3) | 3 (2) | 4 (3) | 4 (3) | 5 (4) | 6 (4) | 0.241 | <0.001 | 0.786 (0.626, 0.946) |
|  | 1-2 | 5 (4) | 6 (4) | 7 (5) | 8 (5) | 8 (5) | 9 (6) |  |  |  |
|  | >2 | 8 (5) | 9 (5) | 9 (5) | 11 (6) | 12 (7) | 12 (7) |  |  |  |
| **MDS-UPDRS III Score – ON** |  |  |  |  |  |  |  |  |  |  |
| Mean (SD) | 0 | 18 (8) | 19 (9) | 18 (9) | 20 (11) | 18 (11) | 20 (14) | 0.412 | <0.001 | 1.504 (1.018, 1.990) |
|  | 1-2 | 22 (9) | 23 (10) | 23 (11) | 23 (12) | 23 (12) | 24 (12) |  |  |  |
|  | >2 | 24 (10) | 24 (11) | 22 (10) | 26 (12) | 26 (13) | 26 (14) |  |  |  |
| **Montreal Cognitive Assessment** |  |  |  |  |  |  |  |  |  |  |
| Mean (SD) | 0 | 27.5 (2.3) | 27.5 (2.5) | 27.5 (2.3) | 27.8 (2.5) | 27.8 (2.2) | 27.4 (2.5) | 0.002 | 0.605 | -0.056 (-0.270, 0.157) |
|  | 1-2 | 27.1 (2.3) | 26.5 (2.7) | 26.5 (3.2) | 26.6 (2.7) | 26.9 (2.9) | 27.0 (2.7) |  | 0.211 | 0.057 (-0.032, 0.146) |
|  | >2 | 26.7 (2.6) | 26.5 (2.9) | 26.3 (3.1) | 26.0 (3.3) | 25.9 (3.9) | 25.7 (4.5) |  | <0.001 | -0.254 (-0.396, -0.111) |
| **Lowest Putamen Ratio** |  |  |  |  |  |  |  |  |  |  |
| Mean (SD) | 0 | 0.36 (0.11) | 0.34 (0.09) | 0.31 (0.08) | N/A | 0.26 (0.07) | N/A | 0.225 | <0.001 | -0.021 (-0.024, -0.017) |
|  | 1-2 | 0.36 (0.12) | 0.32 (0.10) | 0.28 (0.09) | N/A | 0.24 (0.09) | N/A |  |  |  |
|  | >2 | 0.36 (0.13) | 0.30 (0.12) | 0.26 (0.11) | N/A | 0.23 (0.10) | N/A |  |  |  |
| **Mean Striatum** |  |  |  |  |  |  |  |  |  |  |
| Mean (SD) | 0 | 1.57 (0.37) | 1.48 (0.35) | 1.33 (0.28) | N/A | 1.14 (0.25) | N/A | 0.012 | <0.001 | -0.115 (-0.141, -0.090) |
|  | 1-2 | 1.47 (0.40) | 1.28 (0.34) | 1.20 (0.35) | N/A | 1.04 (0.32) | N/A |  | <0.001 | -0.082 (-0.092, -0.072) |
|  | >2 | 1.43 (0.40) | 1.22 (0.38) | 1.10 (0.38) | N/A | 0.99 (0.36) | N/A |  | <0.001 | -0.068 (-0.084, -0.053) |
| **Orthostatic Hypotension (≥ 20 mm Hg change in systolic BP *or* ≥ 10 mm Hg change in diastolic BP)** |  |  |  |  |  |  |  |  |  |  |
| N (%) | 0 | 4 (7%) | 2 (5%) | 3 (10%) | 6 (21%) | 3 (11%) | 1 (4%) | 0.767 | 0.329 | 1.070 (0.934, 1.224)<sup>*</sup> |
|  | 1-2 | 59 (13%) | 36 (12%) | 36 (16%) | 26 (15%) | 24 (14%) | 30 (19%) |  |  |  |
|  | >2 | 63 (18%) | 45 (21%) | 25 (17%) | 28 (23%) | 22 (20%) | 27 (26%) |  |  |  |

Note: Severity is defined as none (0 affected domains), mild (1–2 affected domains), or severe (>2 affected domains).
Descriptive summaries (mean [SD] or n [%]) included 874 participants with PD, categorized by baseline autonomic symptom severity as none (n=61), mild (n=469), severe (n=344).. The longitudinal analysis assessed the longitudinal progression of autonomic symptoms from Year 1 through Year 5 and included 589 participants with PD who had autonomic data at baseline and at least one follow-up visit: none (n=44), mild (n=318), severe (n=227) at baseline.
Significance level for comparisons is p<0.007 (after Bonferroni correction).
\* Corresponds to the average odds ratio (95% CI) for a one-year change across all severities.

#### Non-Motor Symptom Progression

PD participants with more autonomic complaints at baseline exhibited significantly greater worsening in MDS-UPDRS I compared to those with fewer complaints as demonstrated by a significant interaction (p = 0.03). The estimated annual change in MDS-UPDRS I score was 0.499 units (95% CI: 0.201, 0.797; p = 0.001) in the ‘none’ category, 0.742 units (95% CI: 0.566, 0.918; p < 0.001) in the ‘mild’ category, and 1.034 units (95% CI: 0.786, 1.283; p < 0.001) in the ‘severe’ category (Table 3 and Supplementary Figure 1). After adjustment for multiple comparisons, the interaction was no longer significant. Similarly, differences in the progression of MDS-UPDRS Part I were no longer statistically significant after weighting in the IPW-GEE model (Interaction P=0.068; Supplementary Table 7).

#### Motor Symptom Progression

Progression of motor symptoms over time was not different based on number of autonomic symptom complaints for motor activities of daily living (MDS-UPDRS part II; p = 0.241) or motor examination (MDS-UPDRS part III ON score; p = 0.412). The estimated annual change in MDS-UPDRS part II score was 0.786 units (95% CI: 0.626, 0.946; p < 0.001), and the estimated annual change in MDS-UPDRS III ON score was 1.504 units (95% CI: 1.018, 1.990; p < 0.001), suggesting an increase over time regardless of number of autonomic symptoms. The IPW-GEE results were consistent with the primary unweighted analyses, showing no corresponding differences in motor decline among participants with greater baseline autonomic symptoms (Supplementary Table 7).

#### Cognitive Decline

Decline in MoCA was different over time (p = 0.002) depending on number of autonomic symptoms experienced at baseline (Table 3 and Supplementary Figure 1). The estimated annual change in MoCA score did not significantly change over time in either the ‘none’ (p = 0.605) or ‘mild’ (p = 0.211) categories. Only participants reporting ‘severe’ autonomic symptoms at baseline had significantly worsened MoCA over time, with an estimated annual change of -0.254 units (95% CI: -0.396, -0.111; p < 0.001). The IPW-GEE results were consistent, showing a difference in decline in MoCA depending on autonomic severity (interaction p=0.002) and a significant change in MoCA over time only among those with ‘severe’ autonomic symptoms at baseline (p=0.003; Supplementary Table 7).

#### Biomarker Correlations

Reporting more autonomic symptoms did not influence changes in % age-/sex-expected putamen DaT uptake over time (p = 0.225). The estimated annual change in lowest putamen ratio was -0.021 units (95% CI: -0.024, -0.017; p < 0.001), suggesting a decrease over time regardless of autonomic severity group. In contrast, there were differences in decline in striatum DaT uptake between groups (p = 0.012) (Table 3 and Supplementary Figure 1). The estimated annual change in mean striatum striatal binding ratio (SBR) is -0.115 units (95% CI: -0.141, -0.090; p < 0.001) in the ‘none’ category, -0.082 units (95% CI: -0.092, -0.072; p < 0.001) in the ‘mild’ category, and -0.068 units (95% CI: -0.084, -0.053; p < 0.001) in the ‘severe’ category, but after adjustment for multiple comparisons, the interaction was no longer significant.

#### Orthostatic hypotension

Reporting more autonomic symptoms at baseline did not influence changes in orthostatic hypotension over time (p = 0.767). The estimated annual odds ratio for orthostatic hypotension was 1.070 (95% CI: 0.934, 1.224), suggesting no increase over time regardless of the number of autonomic symptoms. Likewise, there was no indication of an increase in orthostatic hypotension over time after weighting in the IPW-GEE model (Supplementary Table 7).

## Discussion

This study, using data from a large cohort of early, de novo Parkinson’s disease participants with positive CSF SAA (PPMI) [13], demonstrates that autonomic symptoms in PD progress steadily over a 60-month period. Gastrointestinal and urinary symptoms were the most prominent and burdensome autonomic disturbances, with a notable increase in gastrointestinal dysfunction observed over time. The reliance on medications, particularly those addressing constipation and urinary issues, increased as the disease advanced, reflecting the escalating severity of autonomic disturbances. The presence of comorbidities-especially cardiovascular, metabolic, and urinary disorders-may further exacerbate autonomic symptoms and contribute to greater symptom severity.

Urinary and gastrointestinal symptoms were the most frequent autonomic symptoms reported by PD participants in this cohort. This may reflect SCOPA-AUT structure as there are more GI/urinary questions compared to some other domains, although our focus was on symptom presence. Most participants experienced persistent urinary symptoms throughout the 60-month observation period, and gastrointestinal dysfunction was similarly prevalent. These findings align with previous research, including a Chinese cohort study that identified dry mouth (59.26%) and constipation (49.07%) as the most common gastrointestinal symptoms, and urinary urgency and nocturia (35%) as prominent urinary complaints[15]. The high prevalence of these symptoms in both Western and Chinese populations underscores their central role in the clinical picture of PD. However, the pattern of autonomic involvement can vary across studies. Some investigations have emphasized cardiovascular dysfunction and thermoregulatory abnormalities as frequent features of autonomic impairment in PD[16]. Differences in patient selection, such as the inclusion of atypical parkinsonism, disease stage (early vs. advanced PD)[16, 17], assessment tools (SCOPA-AUT vs. NMSQuest)[18], and racial or ethnic background may contribute to these discrepancies[1]. Additionally, comorbid conditions such as cardiovascular, metabolic, and urinary disorders, which are more prevalent in PD, may influence both the manifestation and severity of autonomic symptoms (Gu et al., 2023).

The high prevalence of gastrointestinal and urinary dysfunction in PD likely reflects underlying neuropathological processes[16]. Alpha-synuclein deposition within the autonomic nervous system leads to neuronal loss and impaired function[19]. The enteric nervous system (ENS), responsible for gastrointestinal regulation, is particularly vulnerable due to its exposure to environmental toxins and pathogens[20]. Similarly, degeneration of Onuf’s nucleus, which controls the external urethral sphincter, contributes to urinary dysfunction[18]. While alpha-synuclein pathology affects the entire autonomic nervous system, the ENS and Onuf’s nucleus may be especially susceptible due to their unique anatomical and physiological characteristics[19]. Given the burden of these symptoms, clinicians should routinely screen for constipation, urinary urgency, nocturia, and related issues during assessments of individuals at risk for or newly diagnosed with PD[1, 18]. Further research is needed to clarify risk factors, underlying mechanisms, and targeted interventions to alleviate these symptoms and improve patients’ quality of life.

While our study confirms a significant progression of both motor and non-motor symptoms over time in PD, as evidenced by the annual changes in MDS-UPDRS II and III scores, we did not find a statistically significant association between the rate of this progression and the number of autonomic symptom complaints at baseline. This suggests that while autonomic dysfunction and motor symptoms both worsen over time in PD, the severity of autonomic symptoms at baseline does not appear to predict a faster rate of motor decline. This finding contrasts with some prior studies suggesting a link between autonomic dysfunction and faster disease progression[3]. However, it is important to note that autonomic dysfunction is highly prevalent in PD[16], and its presence may broadly contribute to disease severity without necessarily influencing the rate of motor decline. Furthermore, fewer autonomic symptoms were associated with more decline in DaTscan uptake in the striatum, [21, 22], suggesting other drivers of dopaminergic dysfunction[11]. Further, the absence of statistical significance in the interaction term indicates that the relationship between autonomic dysfunction and motor decline is not uniform and may be influenced by genetic, lifestyle, or comorbid factors, as well as specific patterns of autonomic dysfunction[11]. Future research is needed to explore the complex interplay between autonomic and motor systems in PD and to identify potential predictors of disease progression.

The relationship between autonomic dysfunction and cognitive decline in PD is complex[23]. Individuals with more than two affected autonomic domains at baseline experienced a steeper decline in cognitive performance, as measured by the MoCA, whereas those with none or one to two affected domains showed no significant change. Cognitive deterioration in PD is multifactorial, involving dopamine and acetylcholine deficits and the spread of alpha-synuclein pathology to cortical regions[24]. Autonomic dysfunction may indirectly contribute to cognitive decline by disrupting sleep, mood, and overall well-being, all of which are known to affect cognitive function[25]. Since the effect was modest in magnitude in our study, it is best regarded as a group-level signal requiring replication rather than as a clinically actionable prognostic marker.

Despite the PD group having a higher frequency of orthostatic hypotension (OH) and the HC group having a lower frequency, we also found no significant longitudinal differences in the change in presence of OH between PD and HC over 60 months, nor any influence from baseline autonomic symptom severity on the SCOPA-AUT scale, which contrasts with the literature. Methodological differences may explain this. For example, Beach and McKay 2024 [26] and Chen et al. 2021 [27] relied on clinical diagnosis in unfiltered PPMI idiopathic PD cohorts without SAA confirmation; our SAA+ restriction ensured more biologically homogeneous early PD with potentially milder profiles. Our longer follow-up with attrition may bias toward milder cases, compared to shorter studies like Merola et al. 2018 (12-month follow-up) [7] or Stanković et al. 2019 (36-month follow-up) [10]. Extensive adjustments for comorbidities and medications likely attenuated effects, differing from the minimal adjustments in Beach and Mckay 2024 [26]. These methodological gaps also call for larger, multi-site trials with consistent OH protocols and full biomarker integration to clarify its true role in early disease.

This study’s strengths include its longitudinal design and the use of a well-characterized cohort, which enabled the prospective assessment of autonomic dysfunction progression over an extended period. However, several limitations must be acknowledged. Reliance on the SCOPA-AUT questionnaire introduces potential recall bias. The SCOPA-AUT is a screening and severity instrument and is not equivalent to a targeted clinical or laboratory autonomic examination. Therefore, symptoms that a participant does not perceive will not be reported, producing under-ascertainment. Conversely, symptoms may be misinterpreted or misattributed, so that complaints arising from benign prostatic hyperplasia, diabetes, medication effects, or normal aging may be recorded as autonomic in origin, producing over-ascertainment. Because these sources of misclassification apply to both cohorts, they are likely to be largely non-differential and would tend to attenuate rather than generate the between group differences reported here; nonetheless, all prevalence estimates should be read as approximations. Antiparkinsonian medications can influence autonomic function[28]. Although statistical models adjusted for medication classes, disentangling drug effects from disease-related autonomic dysfunction remains challenging. Moreover, adjusting for actual medication dose levels, rather than dichotomized medication classes, would have been an ideal way to account for dynamic changes in medication use over time. Cardiovascular, metabolic, and urinary comorbidities-more common in PD-may confound the observed associations, despite statistical adjustments[11]. Importantly, substantial attrition occurred over the five-year period. This raises concern for attrition bias, as those with more severe autonomic symptoms or greater overall disability may have been disproportionately lost to follow-up., As a result, the observed high prevalence of autonomic symptoms should be interpreted cautiously[29]. Nevertheless, the sensitivity analyses showed that PD participants with and without Year 5 autonomic data did not differ significantly in baseline autonomic symptom severity or in the prevalence in any individual autonomic domain at baseline (Supplementary Table 6), and the IPW-GEE results generally supported the robustness of the primary longitudinal findings (Supplementary Table 7). Since heart rate criteria and supine hypertension were not assessed, the OH screening only reflects orthostatic blood pressure, making it impossible to distinguish between neurogenic and non-neurogenic OH. Lastly, the binary indicator of a total autonomic symptom score >1 is likely influenced by a ceiling effect in the PD cohort (96-98% at every visit) and is also high (87-94% at every visit) among HC participants. This limited variability may reduce the ability to detect differences in longitudinal trajectories between the PD and HC groups. Therefore, these findings should be interpreted with caution as they do not necessarily support specificity of autonomic dysfunction in PD.

In conclusion, these findings highlight the critical importance of monitoring autonomic symptoms in PD, given their prevalence, their impact on quality of life, and, among participants with the greatest baseline burden, their association with faster cognitive decline. Future research should prioritize interventions aimed at mitigating autonomic symptoms and explore the links between specific autonomic disturbances and other non-motor symptoms such as sleep disorders, depression, and anxiety[25]. Personalized approaches to managing autonomic dysfunction, tailored to individual patient profiles, may improve outcomes and quality of life[30].

## Supplemental Files

**Supplementary Figure 1:**
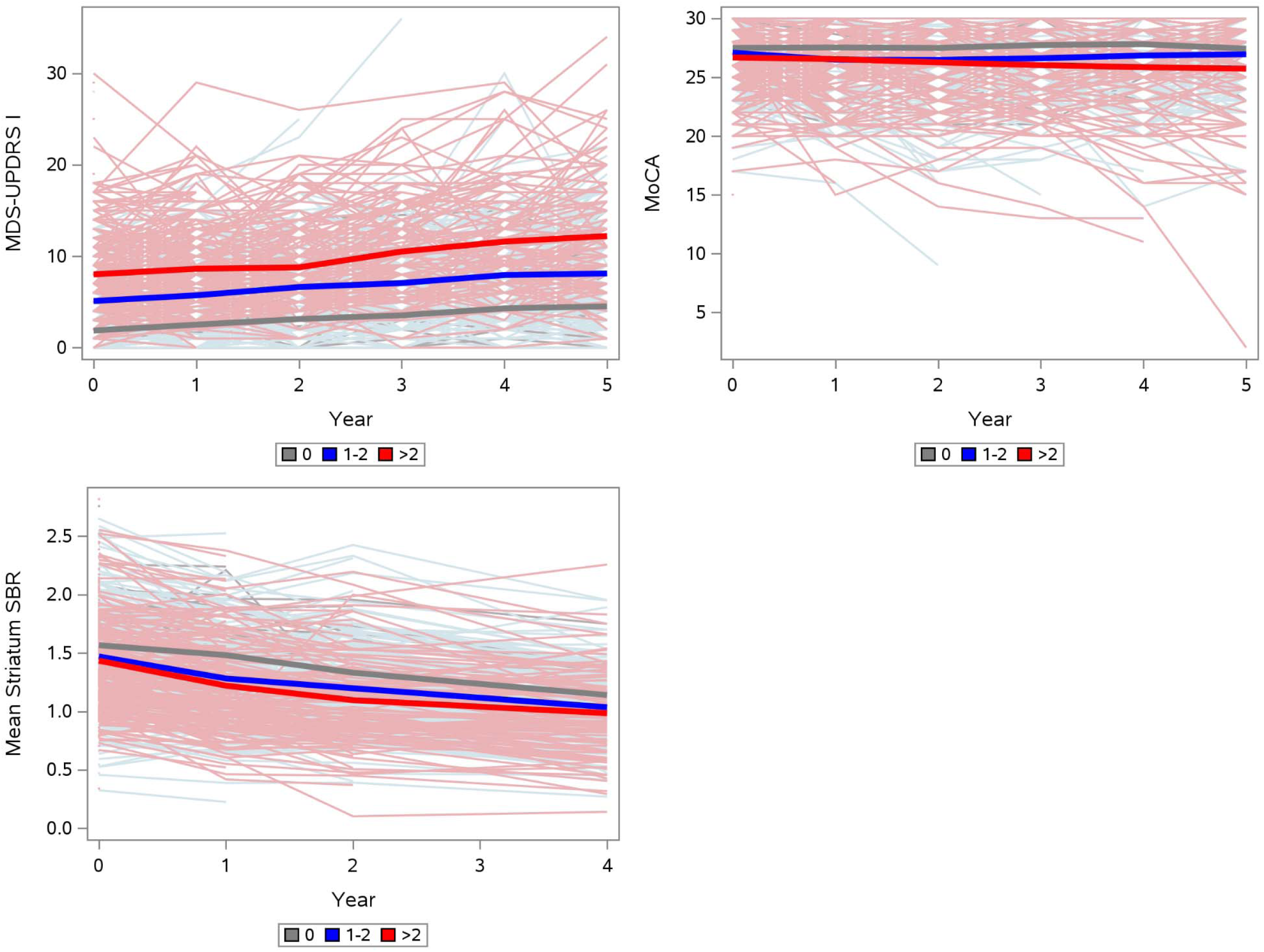
Effect of Autonomic Symptoms at Baseline on Clinical measures Over Time in PD. The PD cohort was stratified into groups based on number of autonomic symptoms reported at baseline (none, 1-2, or >2), with x-axis showing time and y-axis showing MDS-UPDRS Part 1, MoCA, or mean striatum SBR. Bolded lines show means at each visit. MoCA: Montreal Cognitive Assessment; SBR: specific binding ratio.

**Supplementary Table 1:** Frequency of Affected Autonomic Domains.

| **Cohort<br>PD<br>HC** | **# of Affected<br>Autonomic<br>Domains** | **Baseline<br>N=874<br>N=252** | **Month 12<br>N=571<br>N=219** | **Month 24<br>N=408<br>N=182** | **Month 36<br>N=325<br>N=157** | **Month 48<br>N=309<br>N=153** | **Month 60<br>N=287<br>N=145** |
|---|---|---|---|---|---|---|---|
| PD | 0 | 61 (7%) | 33 (6%) | 19 (5%) | 14 (4%) | 11 (4%) | 9 (3%) |
|  | 1 | 196 (22%) | 105 (18%) | 70 (17%) | 54 (17%) | 34 (11%) | 34 (12%) |
|  | 2 | 273 (31%) | 174 (30%) | 124 (30%) | 79 (24%) | 88 (28%) | 75 (26%) |
|  | 3 | 210 (24%) | 153 (27%) | 104 (25%) | 86 (26%) | 90 (29%) | 79 (28%) |
|  | 4 | 93 (11%) | 72 (13%) | 64 (16%) | 58 (18%) | 57 (18%) | 56 (20%) |
|  | 5 | 35 (4%) | 28 (5%) | 19 (5%) | 32 (10%) | 25 (8%) | 26 (9%) |
|  | 6 | 6 (1%) | 6 (1%) | 8 (2%) | 2 (1%) | 4 (1%) | 8 (3%) |
| HC | 0 | 37 (15%) | 41 (19%) | 35 (19%) | 16 (10%) | 19 (12%) | 19 (13%) |
|  | 1 | 114 (45%) | 86 (39%) | 63 (35%) | 63 (40%) | 56 (37%) | 60 (41%) |
|  | 2 | 70 (28%) | 66 (30%) | 57 (31%) | 56 (36%) | 55 (36%) | 41 (28%) |
|  | 3 | 19 (8%) | 17 (8%) | 19 (10%) | 16 (10%) | 13 (8%) | 14 (10%) |
|  | 4 | 10 (4%) | 7 (3%) | 6 (3%) | 4 (3%) | 9 (6%) | 9 (6%) |
|  | 5 | 2 (1%) | 2 (1%) | 2 (1%) | 2 (1%) | 1 (1%) | 2 (1%) |
|  | 6 | 0 (0%) | 0 (0%) | 0 (0%) | 0 (0%) | 0 (0%) | 0 (0%) |

Note: Presence of one affected autonomic domain is defined as a SCOPA-AUT subscore > 1 on one of six domains: gastrointestinal, urinary, cardiovascular, thermoregulatory, pupillomotor, sexual. Autonomic symptom severity was then categorized as none (0 affected domains), mild (1–2 affected domains), or severe (>2 affected domains).

**Supplementary Table 2:** Number of Participants Taking Medications for Autonomic Dysfunction.

|  |  | **Enrolled Participants** |  |  |  |  |  |
|---|---|---|---|---|---|---|---|
| **Cohort**<br>**PD**<br>**HC** | **Medications for Autonomic Dysfunction** | **Baseline**<br>**N=874**<br>**N=252** | **Month 12**<br>**N=571**<br>**N=219** | **Month 24**<br>**N=408**<br>**N=182** | **Month 36**<br>**N=325**<br>**N=157** | **Month 48**<br>**N=309**<br>**N=153** | **Month 60**<br>**N=287**<br>**N=145** |
| PD | Hypotension | 0 (0%) | 1 (<1%) | 2 (<1%) | 4 (1%) | 4 (1%) | 7 (2%) |
|  | Sexual Dysfunction | 87 (10%) | 65 (11%) | 46 (11%) | 44 (14%) | 37 (12%) | 41 (14%) |
|  | Constipation | 149 (17%) | 134 (23%) | 111 (27%) | 97 (30%) | 100 (32%) | 97 (34%) |
|  | Anorexia, Nausea, Vomiting | 0 (0%) | 1 (<1%) | 1 (<1%) | 2 (1%) | 2 (1%) | 2 (1%) |
|  | Drooling | 2 (<1%) | 1 (<1%) | 2 (<1%) | 2 (1%) | 2 (1%) | 2 (1%) |
|  | Urinary Dysfunction | 104 (12%) | 82 (14%) | 64 (16%) | 66 (20%) | 57 (18%) | 54 (19%) |
|  | SCOPA-AUT 26c: Blood Pressure (Hypertension) | 291 (33%) | 194 (34%) | 142 (35%) | 115 (35%) | 110 (36%) | 97 (34%) |
|  | SCOPA-AUT 26d: Other | 283 (32%) | 195 (34%) | 151 (37%) | 120 (37%) | 114 (37%) | 103 (36%) |
| HC | Hypotension | 0 (0%) | 0 (0%) | 0 (0%) | 0 (0%) | 0 (0%) | 0 (0%) |
|  | Sexual Dysfunction | 21 (8%) | 15 (7%) | 15 (8%) | 15 (10%) | 15 (10%) | 10 (7%) |
|  | Constipation | 19 (8%) | 20 (9%) | 18 (10%) | 22 (14%) | 21 (14%) | 20 (14%) |
|  | Anorexia, Nausea, Vomiting | 0 (0%) | 0 (0%) | 0 (0%) | 0 (0%) | 0 (0%) | 0 (0%) |
|  | Drooling | 0 (0%) | 0 (0%) | 0 (0%) | 0 (0%) | 0 (0%) | 0 (0%) |
|  | Urinary Dysfunction | 18 (7%) | 13 (6%) | 13 (7%) | 9 (6%) | 10 (7%) | 10 (7%) |
|  | SCOPA-AUT 26c: Blood Pressure (Hypertension) | 80 (32%) | 79 (36%) | 77 (42%) | 66 (42%) | 62 (41%) | 56 (39%) |
|  | SCOPA-AUT 26d: Other | 81 (32%) | 82 (37%) | 72 (40%) | 59 (38%) | 67 (44%) | 58 (40%) |

Note: Participants may be on more than one medication.
The SCOPA-AUT questions refer to any medication use in the past month.  Other medications come from log records and refer to medication use on the visit date.

**Supplementary Table 3:** Number of Participants with Comorbidities.

|  |  | **Enrolled Participants** |  |  |  |  |  |
|---|---|---|---|---|---|---|---|
| **Cohort**<br>**PD**<br>**HC** | **Comorbidities** | **Baseline**<br>**N=874**<br>**N=252** | **Month 12**<br>**N=571**<br>**N=219** | **Month 24**<br>**N=408**<br>**N=182** | **Month 36**<br>**N=325**<br>**N=157** | **Month 48**<br>**N=309**<br>**N=153** | **Month 60**<br>**N=287**<br>**N=145** |
| PD | Cardiovascular | 335 (38%) | 236 (41%) | 184 (45%) | 159 (49%) | 152 (49%) | 151 (53%) |
|  | Metabolic | 94 (11%) | 62 (11%) | 54 (13%) | 54 (17%) | 52 (17%) | 52 (18%) |
|  | Urinary | 93 (11%) | 85 (15%) | 66 (16%) | 58 (18%) | 59 (19%) | 55 (19%) |
| HC | Cardiovascular | 100 (40%) | 92 (42%) | 88 (48%) | 75 (48%) | 74 (48%) | 69 (48%) |
|  | Metabolic | 30 (12%) | 28 (13%) | 26 (14%) | 21 (13%) | 23 (15%) | 24 (17%) |
|  | Urinary | 17 (7%) | 17 (8%) | 16 (9%) | 10 (6%) | 13 (8%) | 15 (10%) |

Note: Participants may have more than one comorbidity.

**Supplementary Table 4:** Model Adjustments for Table 2.

| Label | Baseline Characteristics | Classes of Medication | Comorbidities |
|---|---|---|---|
| SCOPA AUT Total Score (>1) | Age, Sex, Baseline Response | Sexual Dysfunction, Constipation, Urinary Dysfunction, Hypertension, Other | Cardiovascular, Metabolic, Urinary |
| SCOPA AUT Gastrointestinal Dysfunction (>1) | Age, Sex, Baseline Response | Constipation, ‘Anorexia, Nausea, Vomiting’ |  |
| SCOPA AUT Urinary Dysfunction (>1) | Age, Sex, Baseline Response | Urinary Dysfunction | Urinary |
| SCOPA AUT Cardiovascular Dysfunction (>1) | Age, Sex, Baseline Response | Hypotension, Hypertension | Cardiovascular, Metabolic |
| SCOPA AUT Thermoregulatory Dysfunction (>1) | Age, Sex, Baseline Response | Hypotension, Hypertension, ‘Anorexia, Nausea, Vomiting’ |  |
| SCOPA AUT Pupillomotor Dysfunction (>1) | Age, Sex, Baseline Response | Hypotension, Hypertension |  |
| SCOPA AUT Sexual Dysfunction (>1) | Age, Sex, Baseline Response | Sexual Dysfunction |  |
| SCOPA AUT Sexual Dysfunction Men (>1) | Age, Baseline Response | Sexual Dysfunction |  |
| SCOPA AUT Sexual Dysfunction Women (>1) | Age, Baseline Response |  |  |
| Orthostatic Hypotension | Age, Sex, Baseline Response | Hypotension, Hypertension | Cardiovascular, Metabolic |

**Supplementary Table 5:**
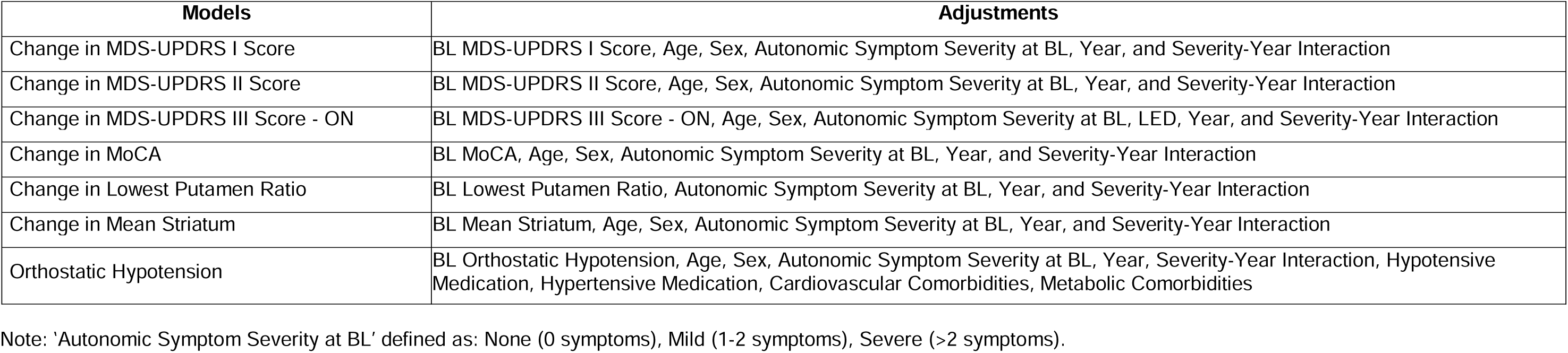
Model Adjustments for Table 3.

| **Models** | **Adjustments** |
|---|---|
| Change in MDS-UPDRS I Score | BL MDS-UPDRS I Score, Age, Sex, Autonomic Symptom Severity at BL, Year, and Severity-Year Interaction |
| Change in MDS-UPDRS II Score | BL MDS-UPDRS II Score, Age, Sex, Autonomic Symptom Severity at BL, Year, and Severity-Year Interaction |
| Change in MDS-UPDRS III Score - ON | BL MDS-UPDRS III Score - ON, Age, Sex, Autonomic Symptom Severity at BL, LED, Year, and Severity-Year Interaction |
| Change in MoCA | BL MoCA, Age, Sex, Autonomic Symptom Severity at BL, Year, and Severity-Year Interaction |
| Change in Lowest Putamen Ratio | BL Lowest Putamen Ratio, Autonomic Symptom Severity at BL, Year, and Severity-Year Interaction |
| Change in Mean Striatum | BL Mean Striatum, Age, Sex, Autonomic Symptom Severity at BL, Year, and Severity-Year Interaction |
| Orthostatic Hypotension | BL Orthostatic Hypotension, Age, Sex, Autonomic Symptom Severity at BL, Year, Severity-Year Interaction, Hypotensive Medication, Hypertensive Medication, Cardiovascular Comorbidities, Metabolic Comorbidities |

Note: ‘Autonomic Symptom Severity at BL’ defined as: None (0 symptoms), Mild (1-2 symptoms), Severe (>2 symptoms).

**Supplementary Table 6:**
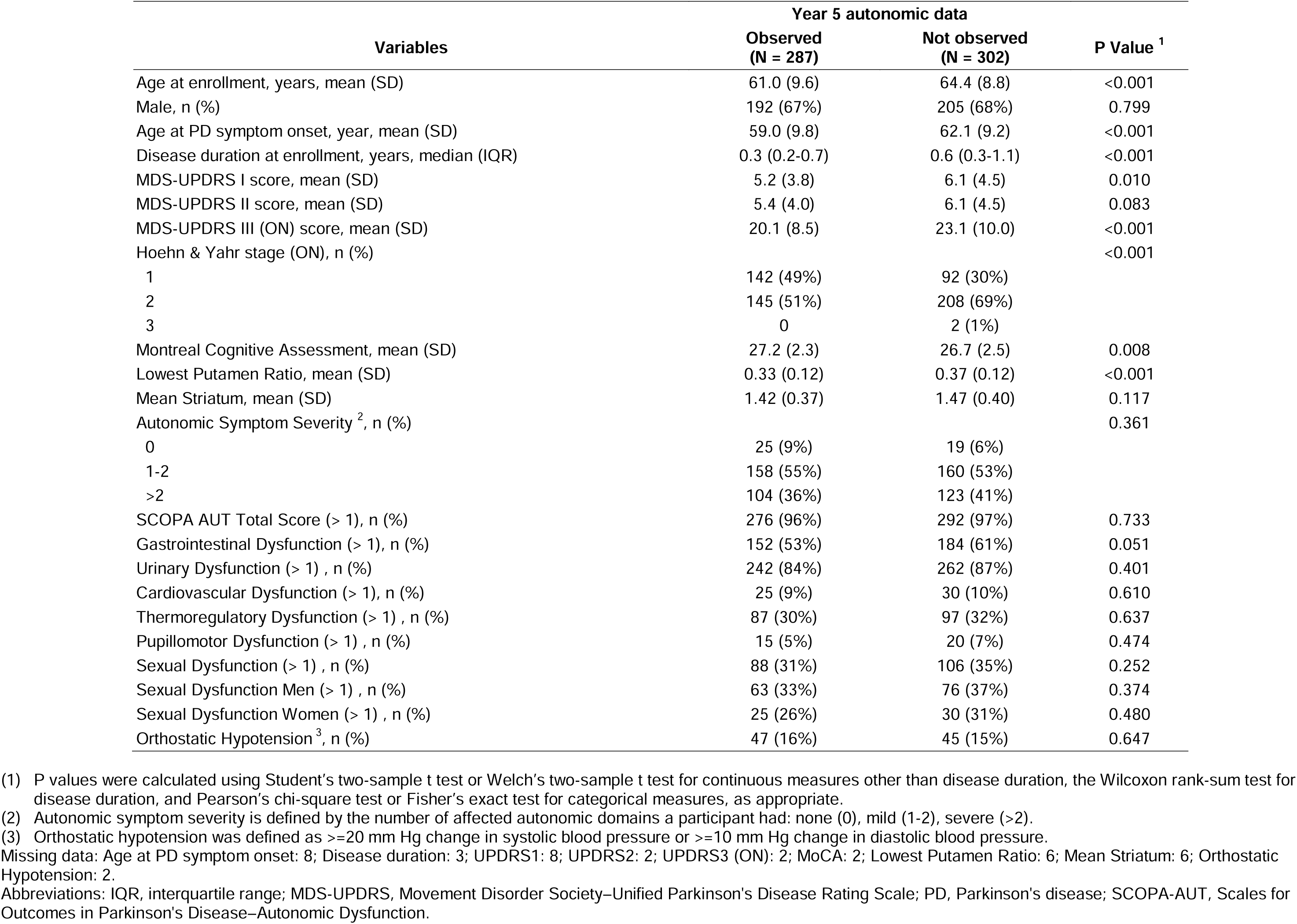
Baseline Characteristics of Participants with PD Who Had at Least One Follow-up Visit, by Year 5 Autonomic Data Availability.

**Supplementary Table 7.** IPW-GEE Sensitivity Analysis of Clinical Outcomes Over Time by Baseline Autonomic Symptom Severity in Participants with PD.

| Clinical Outcome | Severity<br>None (0)<br>Mild (1-2)<br>Severe (>2) | p-value<br>Interaction<br>(Group x Time) | p-value<br>Time | Time Effect<sup>4</sup> (95% CI) |
|---|---|---|---|---|
| MDS-UPDRS I Score | Overall | 0.068 | <.001 | 0.722 (0.566, 0.879) |
| MDS-UPDRS II Score | Overall | 0.501 | <.001 | 0.816 (0.625, 1.006) |
| MDS-UPDRS III (ON) Score | Overall | 0.480 | <.001 | 1.484 (0.925, 2.042) |
| Montreal Cognitive Assessment | Overall | 0.002 |  |  |
|  | None (0) |  | 0.449 | -0.097 (-0.349, 0.154) |
|  | Mild (1-2) |  | 0.071 | 0.088 (-0.007, 0.183) |
|  | Severe (>2) |  | 0.003 | -0.232 (-0.385, -0.079) |
| Orthostatic Hypotension<sup>5</sup> | Overall | 0.390 | 0.823 | 1.017 (0.879, 1.176) |

(1) Sensitivity analyses of longitudinal changes in clinical outcomes from Year 1 through Year 5 were performed using inverse probability-weighted generalized estimating equation (IPW-GEE) models. The analytic population included participants with Parkinson’s disease (PD) who had baseline autonomic data and autonomic data from at least one follow-up visit. IPWs were calculated as the inverse of the estimated cumulative probability of having observed autonomic data at each follow-up visit. These probabilities were estimated using visit-specific logistic regression models of the probability of being observed at a given visit conditional on baseline characteristics (age, sex, disease duration, and lowest putamen ratio) and prior-visit characteristics (MDS-UPDRS Parts I, II, and III [ON] scores; Montreal Cognitive Assessment [MoCA] score; and changes in systolic and diastolic blood pressure).
(2) Clinical outcomes included in the IPW and IPW-GEE models *(*MDS-UPDRS Parts I, II, III (ON) scores, MoCA, changes in systolic and diastolic blood pressure, and orthostatic hypotension) with intermittent missing values were replaced using linear interpolation. Values before the first or after the last observation visit were not imputed.
(3) Baseline autonomic symptom severity was classified as none (0 symptoms), mild (1-2 symptoms), or severe (>2 symptoms).
(4) For continuous outcomes (MDS-UPDRS Parts I, II, and III [ON] scores; Montreal Cognitive Assessment [MoCA] score), the time effect represents the estimated mean change in the outcome per year, with a 95% confidence interval (CI). For binary outcomes (orthostatic hypotension), the time effect is expressed as the odds ratio (OR) for the presence of orthostatic hypotension associated with each additional year of follow-up, with a 95% CI; an OR greater than 1 indicates increasing odds over time.
(5) Orthostatic hypotension was defined as >=20 mm Hg change in systolic blood pressure or >=10 mm Hg change in diastolic blood pressure.
Note: Significance level for comparisons is p<0.01 (after Bonferroni correction).
Abbreviations: PD, Parkinson’s disease; CI, confidence interval; GEE, generalized estimating equations; IPW, inverse probability weighting.

Supplementary Author List for Parkinson’s Progression Markers Initiative authors

## Data Availability

Data used in the preparation of this article were obtained on January 8, 2024 from the Parkinson Progression Markers Initiative (PPMI) database (https://www.ppmi-info.org/access-data-specimens/download-data), RRID:SCR_006431. For up-to-date information on the study, visit https://www.ppmi-info.org. This analysis was conducted by the PPMI Statistics Core and used actual dates of activity for participants, a restricted data element not available to public users of PPMI data. Statistical analysis codes used to perform the analyses in this article are shared on Zenodo (https://doi.org/10.5281/zenodo.15739155).

## PPMI STUDY COMMITTEES, CORES, AND COLLABORATORS

### PPMI EXECUTIVE STEERING COMMITTEE

Kenneth Marek, MD^1^ (Principal Investigator); Tanya Simuni, MD^2^; Andrew Siderowf, MD^3^; Caroline M. Tanner, MD^4^; Thomas F. Tropea, DO^1^; Tatiana Foroud, PhD^5^; Lana M. Chahine, MD^4^; Brit Mollenhauer, MD^6^; Kalpana Merchant, MD^2^; Douglas Galasko, MD^7^; Christopher S. Coffey, PhD^8^; Kathleen L. Poston, MD^9^; Roseanne D. Dobkin, PhD^10^; Ethan G. Brown, MD^4^; Roy N. Alcalay, MD^11^; Dan Weintraub, MD^3^; Emily Flagg, BA^1^; Kimberly Fabrizio, BA^1^; Craig Stanley, PhD^1^; Bridget A. McMahon, BS^1^

### PPMI STEERING COMMITTEE

Susan Bressman, MD^12^; Cornelis Blauwendraat, PhD^13^; Sonya Dumanis, PhD^13^; Paola Casalin, PhD^14^; Raymond James, RN^15^; Karl Kieburtz, MD^16^; Sneha Mantri, MS^17^; Werner Poewe, MD^18^; Michael Schwarzschild, MD^19^; John Seibyl^1^, MD; David Standaert, PhD^20^; Duygu Tosun-Turgut, PhD^4^

### MICHAEL J. FOX FOUNDATION

Sohini Chowdhury, MA^21^; Jamie Eberling, PhD^21^; Mark Frasier, PhD^21^; Leslie Kirsch, EdD^21^; Katie Kopil, PhD^21^; Maggie Kuhl, BA^21^; Alyssa O’Grady, BA^21^; Todd Sherer, PhD^21^; Tawny Willson, MBS^21^

### PPMI STUDY CORES

*Project Management Core:* Emily Flagg, BA^1^

*Site Management Core:* Thomas F Tropea, DO^1^; Bridget McMahon, BS^1^

*Data Strategy and Technical Operations:* Craig Stanley, PhD^1^; Kim Fabrizio, BA^1^

*Data Management Core:* Dixie Ecklund, MBA^8^, MSN

*Screening Core:* Tatiana Foroud, PhD^5^; Laura Heathers, BA^5^; Christopher Hobbick, BSCE^5^; Gena Antonopoulos, BSN^5^

*Imaging Core:* John Seibyl, MD^1^; Kathleen Poston, MD^9^

*Statistics Core:* Christopher Coffey, PhD ^9^; Chelsea Caspell-Garcia, MS ^9^; Michael Brumm, MS ^9^

*Bioinformatics Core:* Arthur Toga, PhD^22^; Karen Crawford, MLIS^22^

*Biorepository Core:* Tatiana Foroud, PhD^5^; Abigail Erickson, BS^5^

*Biologics Review Committee:* Brit Mollenhauer, MD^6^; Doug Galasko, MD^7^; Kalpana Merchant, MD^2^

*Genetics Core:* Andrew Singleton, PhD^24^

*Pathology Core:* Tatiana Foroud, PhD^5^; Dirk Keene, MD^5^

*Found:* Caroline Tanner, MD^4^; Ethan Brown, MD^4^

*PPMI Online:* Carlie Tanner, MD^4^; Ethan Brown, MD^4^; Lana Chahine, MD^4^; Roseann Dobkin, PhD^10^; Monica Korell, MPH^4^

### PPMI SITE INVESTIGATORS

Neha Prakash MD^1^; Tanya Simuni, MD^2^; Nabila Dahodwala MD^3^; Caroline Tanner, MD^4^; Brit Mollenhauer MD^7^; Susana Schnell MD^6^; Douglas Galasko, MD^7^; Anat Mirelman PhD^11^; Roy Alcalay MD^11^; Katherine Leaver MD^12^; Marie Saint-Hilaire MD^15^; Ruth Schneider MD^16^; Christopher Tarolli MD^16^; Werner Poewe, MD^18^; Corinne Horlings, MD^18^; Aleksandar Videnovic MD^19^; David Standaert PhD^20^; Marissa Dean, MD^20^; Abigail Olsen MD^24^; Rejko Krueger MD^25^; Stewart Factor DO^26^; Joseph Ǫuinn MD^26^; Robert Hauser MD^28^; Amy Amara PhD^29^; Michelle Fullard MD^29^; Cyrus Zabetian MD^30^; Hubert Fernandez MD^31^; Kathrin Brockmann MD^32^; Isabel Wurster PhD^32^; Yen Tai PhD^33^; Paolo Barone PhD^34^; Marina Picillo MD^34^; Stuart Isaacson MD^35^; Alberto Espay MD^36^; Eduardo Tolosa PhD^37^; Alicia Garrido MD^37^; Javier Ruiz Martinez PhD^38^; Leonidas Stefanis PhD^39^; Christos Koros MD^39^; Kelvin Chou MD^40^; Lorraine Kalia MD^41^; Connie Marras PhD^41^; David Grimes MD^42^; Tiago Mestre PhD^42^; Rajesh Pahwa MD^43^; Mark Lew MD^44^; Holly Shill MD^45^; Shyamal Mehta MD^46^; Giulietta Riboldi MD^47^; Nikolaus McFarland PhD^48^; Ron Postuma MD^49^; Zoltan Mari MD^50^; David Ledingham MD^51^; Nicola Pavese PhD^51^; Michele Hu PhD^52^; Norbert Brüggemann MD^53^; Christine Klein MD^53^; Bastiaan Bloem PhD^54^; Cristina Simonet PhD^55^; Alastair Noyce PhD^55^; Anette Janzen PhD^56^; Wolfgang Oertel PhD^56^; Njideka Okubadejo MD^57^; David Shprecher DO^58^; Arjun Tarakad MD^59^; Emile Moukheiber MD^60^

### PPMI SITE COORDINATORS

Joy Anatala¹; Robert Moss¹; Mackenzee George¹; Kristen Hagwell¹; Ashley Romano¹; Sarah Chatfield¹; Stephanie Roman¹; Carla Aranda²; Karen Williams²; Sophia Melton²; Karina Benson²; Sarah Rogers^2^; Ritikha Vengadesh^3^; Joana Salievska^3^; Adaeze Njoku^3^; Taylor Siegal^3^; Jaime Heller⁴; Primi Ranola⁴; Farah Kausar^4^; Jacque Perkins^4^; Diana Willeke^6^; Elizabeth Kalinkara-Gomez^6^; Susanne Scaglione^6^; Janelle Rodriguez^7^; Nobuko Kemmotsu^7^; May Eshel^11^; Deborah Raymond^12^; Emma Margolis^12^; Ivy Gu^12^; Maya Rawal^12^; Abigail Desrosiers^15^; Raymond James^15^; Lauren Jackson^16^; Brittany Clark^16^; Iris Egner^18^; Nathan Fogarty^19^; Courtney Blair^20^; Lauren Ruffrage^20^; Sherri Mosovsky^24^; Berenice Sevilla²⁵; Sonja Jonsdottir^25^; Claire Pauly^25^; Emna Bouhajja^25^; Barbara Sommerfeld²⁶; Madison Christopher²⁷; Montana Pearson²⁸; Erica Botting²⁸; Gabriella Mazur²⁸; Danielle Derlein²⁹; Evan Doll²⁹; Ying Liu²⁹; Nathan Cheng^29^; Laura Gorla^29^; Lucas Lattanzio^29^, Sabrina Akhter Mim^29^; Dillan Patel^29^; Ciera Cobb³⁰; Gianna Dal Porto³⁰; Jennifer Mule³¹; Stuart Houltham³¹; Renee Dancie³¹; Ella Hilt³²; Aldazier Jakiran^33^; Syeda Visha Fatima^33^; Dominga Valentino^34^; Lisbeth Pennente^35^; Bobbie Stubbeman³⁶; Ayesha Shaik^36^; Katherin Galvez^36^; Valeria Ravasi³⁷; Maria Fernanda Farfan Ale³⁷; Ioana Croitoru^38^; Nikolas Papagiannakis³⁹; Evangelia Stanitsa³⁹; Frank Ferrari⁴⁰; Mengyu Zheng⁴¹; Shawna Reddie⁴²; Alicia Alejandra⁴³; Sydney Chappell^43^; Carsyn Smith^43^; Colette Worthington^43^; Alejandra Valenzuela^44^; Aliana Valenzuela^44^; Bethzaida Herrera^45^; Martin Salamanca Jr.^45^; Sara Dresler⁴⁶; Neil Santos^46^; Fahrial Esha⁴⁷; Danayeat Abraha⁴⁷; Mark Belio⁴⁷; Kyle Rizer⁴⁸; Nadine Zablith⁴⁹; Roxana Garcia^50^; Liliana Dumitrescu⁵⁰; Victoria Kate Foster⁵¹; Debra Galley⁵¹; Robyn Iredale⁵¹; Jamil Razzaque⁵²; Madita Grümmer⁵³; Lian Feenstra⁵⁴; Natalie Donkor⁵⁵; Ayat Azhar⁵⁵; Rhiannon Laban⁵⁵; Ruby Lathey⁵⁵; Anisa Shahid⁵⁵; Elisabeth Sittig⁵⁶; Oluwadamilola Ojo⁵⁷; Kelly Clark⁵⁸; Rory Mahabir⁵⁹; Giselle Silva^59^; Ramya Palani⁶⁰; Shamera Willoughby⁶⁰

### INSTITUTIONS AND AFFILIATIONS

1. Institute for Neurodegenerative Disorders; New Haven, CT, USA
2. Northwestern University; Chicago, IL, USA
3. University of Pennsylvania; Philadelphia, PA, USA
4. University of California, San Francisco; San Francisco, CA, USA
5. Indiana University; Indianapolis, IN, USA
6. Paracelsus-Elena Klinik; Kassel, Germany
7. University of California, San Diego, San Diego, CA, USA
8. University of Iowa; Iowa City, IA, USA
9. Stanford University; Stanford, CA, USA
10. Rutgers University; New Brunswick, NJ, USA
11. Tel Aviv Sourasky Medical Center; Tel Aviv, Israel
12. Mount Sinai Beth Israel; New York, NY, USA
13. Coalition for Aligning Science; Chevy Chase, MD, USA
14. BioRep; Milan, Italy
15. Boston University School of Medicine; Boston, MA, USA
16. University of Rochester; Rochester, NY, USA
17. Duke University; Durham, NC, USA
18. University of Innsbruck; Innsbruck, Austria
19. Massachusetts General Hospital; Boston, MA, USA
20. University of Alabama at Birmingham; Birmingham, AL, USA
21. 21. The Michael J. Fox Foundation for Parkinson’s Research; New York, NY, USA
22. Laboratory of Neuroimaging (LONI), USC; Los Angeles, CA, USA
23. National Institute on Aging, NIH; Bethesda, MD, USA
24. University of Pittsburgh; Pittsburgh, PA, USA
25. University of Luxembourg; Esch-sur-Alzette, Luxembourg
26. Emory University; Atlanta, GA, USA
27. Oregon Health and Science University; Portland, OR, USA
28. University of South Florida; Tampa, FL, USA
29. University of Colorado; Aurora, CO, USA
30. VA Puget Sound Health System; Seattle, WA, USA
31. Cleveland Clinic; Cleveland, OH, USA
32. University of Tubingen; Tubingen, Germany
33. Imperial College of London; London, UK
34. University of Salerno; Salerno, Italy
35. Parkinson’s Disease and Movement Disorders Center; Boca Raton, FL, USA
36. University of Cincinnati; Cincinnati, OH, USA
37. Hospital Clinic of Barcelona; Barcelona, Spain
38. Hospital Universitario Donostia; San Sebastian, Spain
39. University of Athens; Athens, Greece
40. University of Michigan; Ann Arbor, MI, USA
41. Toronto Western Hospital; Toronto, Canada
42. The Ottawa Hospital; Ottawa, Canada
43. University of Kansas Medical Center; Kansas City, KS, USA
44. Keck School of Medicine of the University of Southern California; Los Angeles, CA, USA
45. Barrow Neurological Institute; Phoenix, AZ, USA
46. Mayo Clinic Arizona; Scottsdale, AZ, USA
47. NYU Langone Medical Center; New York, NY, USA
48. University of Florida; Gainesville, FL, USA
49. Montreal Neurological Institute and Hospital/McGill; Montreal, ǪC, Canada
50. Cleveland Clinic-Las Vegas Lou Ruvo Center for Brain Health; Las Vegas, NV, USA
51. Clinical Ageing Research Unit; Newcastle, UK
52. John Radcliffe Hospital Oxford and Oxford University; Oxford, UK
53. University of Luebeck; Luebeck, Germany
54. Radboud University; Nijmegen, Netherlands
55. Ǫueen Mary University of London; London, UK
56. Philipps-University Marburg; Marburg, Germany
57. University of Lagos; Lagos, Nigeria
58. Banner Sun Health Research Institute; Sun City, AZ, USA
59. Baylor College of Medicine; Houston, TX, USA
60. Johns Hopkins University; Baltimore, MD, USA

### PPMI STUDY FUNDING PARTNER STATEMENT

PPMI – a public-private partnership – is funded by the Michael J. Fox Foundation for Parkinson’s Research and funding partners, including AbbVie, Alamar Biosciences, Aligning Science Across Parkinson’s (ASAP), Arrowhead Pharma, Arvinas, AskBio, BIAL, BioArctic, Biohaven, BlueRock Therapeutics, Bristol Myers Squibb, Calico Labs, Capsida Biotherapeutics, Critical Path Institute, DaCapo Brainscience, Denali, Edmond J. Safra Foundation, Eli Lilly, Gain Therapeutics, GE Healthcare, Genentech, GSK, Insitro, Johnson C Johnson Innovative Medicine, Lundbeck, Merck, Neumora, Neuron23, Novartis, Olink, Regeneron, Roche, Sanofi, Tenvie, UCB, Vanqua Bio, Voyager Therapeutics, The Weston Family Foundation.

